# Person-centered care competence and patient safety competence in relation to patient safety culture: Mediating effects of patient safety management activities among nurses in central Vietnam

**DOI:** 10.64898/2026.03.09.26347979

**Authors:** Ho Duy Binh, Dang Thi Thanh Phuc, Vo Thi Nhi, Ho Thi My Yen, Nguyen Binh Thao Nguyen, Tran Thi Kim Quyen, Duong Thi Ngoc Lan

**Affiliations:** University of Medicine and Pharmacy, Hue University, Hue, Vietnam; The University of Danang - School of Medicine and Pharmacy, Da Nang, Vietnam

**Keywords:** person-centered care, patient safety competence, patient safety management activities, patient safety culture, nurses, Vietnam, mediation analysis

## Abstract

**Background:** Patient safety culture is a key determinant of healthcare quality, yet evidence remains limited on how nurses’ person-centered care competence and patient safety competence relate to patient safety culture through patient safety management activities, particularly in low- and middle-income settings.

**Objective:** To describe levels of person-centered care competence, patient safety competence, patient safety management activities, and patient safety culture among nurses in central Vietnam, and to examine direct and indirect relationships among these factors using a mediation model.

**Methods:** A multicenter cross-sectional study was conducted among 1,036 nurses from five tertiary hospitals in central Vietnam (response rate 99.6%). Person-centered care competence was measured using P-CAT, patient safety competence using H-PEPSS, patient safety management activities using PSMA, and patient safety culture using HSOPSC. Descriptive statistics, Pearson correlations, and multivariable linear regression analyses were performed. Mediation effects were tested using bias-corrected bootstrapping with 5,000 resamples.

**Results:** Mean scores indicated moderate-to-high levels of person-centered care competence (3.77±0.36), patient safety competence (4.23±0.36), and patient safety management activities (4.44±0.35), while patient safety culture was moderate (3.93±0.35). All variables were positively correlated, with the strongest association observed between person-centered care competence and patient safety culture (r=0.49, p<0.001). In adjusted regression analyses, person-centered care competence and patient safety competence were independently associated with patient safety management activities (β=0.149 and β=0.274; both p<0.001). Patient safety management activities were significantly associated with patient safety culture (β=0.102, p<0.001). Bootstrapped mediation analyses demonstrated significant partial mediation through patient safety management activities for both person-centered care competence (indirect β=0.0265, 95% confidence interval 0.0143-0.0404) and patient safety competence (indirect β=0.0497, 95% confidence interval 0.0304-0.0711).

**Conclusions:** Higher person-centered care competence and patient safety competence were associated with stronger patient safety culture, partly through increased engagement in patient safety management activities. Interventions to strengthen patient safety culture should combine competence development with organizational supports that enable reliable frontline safety practices.

## Introduction

Patient safety culture (PSC) is a critical determinant of healthcare quality and patient outcomes. Strong PSC has been consistently associated with reduced adverse events, improved clinical performance, enhanced organizational learning, and greater patient trust [1], [2], [3], [4], [5]. Nurses play a central role in patient safety because of their continuous presence at the bedside and their responsibility for coordinating complex clinical care processes across shifts and disciplines.

Person-centered care (PCC) is increasingly recognized as a core component of high-quality and safe healthcare delivery. Contemporary evidence indicates that PCC contributes to safer care by improving communication, patient engagement, and shared decision-making, thereby reducing preventable harm [6], [7], [8]. Competence in delivering PCC among nurses has been associated with better adherence to safety practices and improved patient experiences in recent international studies.

Patient safety competence (PSCp), encompassing knowledge, skills, and attitudes related to teamwork, communication, risk management, and human factors, is essential for preventing adverse events. Recent nursing research has shown that higher PSCp is associated with improved safety behaviors and safety outcomes in hospital settings [9], [10], [11]. These findings support the view that competence alone is insufficient unless translated into routine safety practices.

Patient safety management activities (PSMA) represent observable safety behaviors performed by nurses in daily practice, including patient identification, medication safety, infection prevention, and fall prevention. Behavioral and organizational safety theories propose that individual competencies influence safety-related behaviors, which subsequently shape collective safety climate and culture [12], [13]. Recent empirical studies in healthcare settings have further supported the role of frontline safety behaviors as proximal determinants of PSC.

Although previous studies have examined individual determinants of PSC, integrated empirical models incorporating PCC competence, PSCp, and PSMA within a unified mediation framework remain limited, particularly in low- and middle-income countries. Strengthening frontline safety behaviors has been identified as a global priority in international patient safety initiatives, yet evidence from low- and middle-income country nursing contexts remains scarce [4], [5]. Context-specific evidence is therefore needed to inform nursing education, management strategies, and policy interventions.

Accordingly, this study was guided by a theory-informed conceptual framework that integrates individual competencies, frontline safety behaviors, and organizational PSC. This framework draws on three complementary theoretical perspectives. First, the Theory of Planned Behavior posits that individual knowledge, attitudes, and perceived behavioral control shape the likelihood of behavioral enactment, providing a foundation for linking professional competence to observable safety practices [12]. Second, safety climate and safety performance models conceptualize frontline safety behaviors as proximal determinants through which individual characteristics influence collective safety outcomes [13]. Third, Donabedian’s structure–process–outcome model frames professional competence as a structural attribute that affects care processes and, ultimately, organizational outcomes such as PSC [14].

Within this integrated framework, nurses’ PCC competence and PSCp are conceptualized as distal individual determinants. These competencies are expected to influence the consistent enactment of frontline PSMA, which represent proximal safety behaviors embedded in daily nursing practice. Through repeated and reliable performance of PSMA, shared perceptions, norms, and expectations related to patient safety are reinforced, contributing to the development and maintenance of organizational PSC. This pathway is consistent with systems-based patient safety theory, which emphasizes that individual competence must be translated into reliable processes of care to reduce harm in complex healthcare systems [15]. Thus, PSMA is positioned as a theoretically grounded behavioral mediator rather than a purely statistical construct. Therefore, this study aimed to (1) describe the levels of PCC competence, PSCp, PSMA, and PSC among nurses in central Vietnam, and (2) examine direct and indirect relationships among these factors using a mediation model in which person-centered care competence and PSCp influence PSC both directly and indirectly through PSMA.

## Materials and methods

### Study design and reporting guideline

A multicenter cross-sectional study was conducted. This manuscript was prepared in accordance with the Strengthening the Reporting of Observational Studies in Epidemiology (STROBE) guidelines for cross-sectional studies [16].

### Study setting and participants

The study was conducted among nurses working in clinical departments of five tertiary (Grade I) hospitals in central Vietnam. To ensure institutional anonymity and comply with journal requirements, the names of participating hospitals are not disclosed. Eligible participants were registered nurses providing direct patient care and having at least six months of clinical experience at their current workplace. Nurses in administrative positions without routine clinical duties were excluded.

### Sample size determination

Sample size was calculated using G*Power version 3.1 for a correlational study design. The calculation assumed a significance level of α = 0.05, statistical power (1 − β) = 0.8, and a small effect size (f² = 0.02), consistent with recommendations for behavioral and health services research. The minimum required sample size was 945. To account for potential non-response, 10% was added, resulting in a target sample size of 1,040 nurses [17]. A total of 1,036 valid questionnaires were included in the final analysis.

### Sampling procedure

A stratified random sampling method with proportional allocation was employed. Hospitals were first selected using convenience sampling. Within each hospital, the number of nurses invited to participate was determined proportionally based on the total number of eligible nurses using Cochran’s formula for sample allocation in stratified sampling designs [18]. Within each clinical department, nurses were selected using systematic random sampling from staff lists provided by the nursing departments.

### Data collection procedures

Data collection was carried out from June 2024 to December 2024 in collaboration with the nursing departments of participating hospitals. Surveys were administered in paper format during two to three visits to each clinical department to ensure inclusion of nurses across different shifts. Participants were informed about the study objectives, procedures, voluntary nature of participation, and confidentiality measures. Written informed consent was obtained prior to participation. Each questionnaire required approximately 15–20 minutes to complete. Completed questionnaires were reviewed on-site to minimize missing data.

### Measurement instruments

Instruments not previously available in Vietnamese were translated and culturally adapted following the World Health Organization (WHO) process of translation and adaptation of instruments, including forward translation, expert panel review, back-translation, pretesting, and cognitive interviewing [19], [20].

#### Person-centered care competence

PCC competence was measured using the Person-Centred Care Assessment Tool (P-CAT), originally developed by Edvardsson et al. The instrument consists of 13 items across three domains: personalized care, organizational support, and environmental accessibility. Items are rated on a 5-point Likert scale (1 = strongly disagree to 5 = strongly agree), with five negatively worded items reverse-coded. Total scores range from 13 to 65, with higher scores indicating greater PCC competence [21]. In the present study, Cronbach’s alpha for the P-CAT was 0.662 (pilot study: 0.772). Although slightly below the conventional 0.70 threshold, this level is considered acceptable in cross-cultural adaptation studies and exploratory research contexts.

#### Patient safety competence

PSCp was assessed using the Health Professional Education in Patient Safety Survey (H-PEPSS), developed by Ginsburg et al. The H-PEPSS comprises 23 items across six domains, including teamwork, communication, risk management, human and environmental factors, recognition of adverse events, and safety culture. Responses are rated on a 5-point Likert scale (1 = strongly disagree to 5 = strongly agree), with higher scores indicating greater PSCp [9]. Cronbach’s alpha in this study was 0.898 (pilot study: 0.918).

#### Patient safety management activities

PSMA were measured using a 24-item PSMA scale, originally developed and validated among hospital nurses by Park and Kim [22]. The scale covers six domains: patient identification, communication, high-alert medication management, surgical and procedural verification, infection prevention, and fall prevention. Items are rated on a 5-point Likert scale (1 = never to 5 = always), with higher scores indicating more frequent engagement in safety management activities. In the present study, the PSMA demonstrated good internal consistency, with a Cronbach’s alpha of 0.863 (pilot study: 0.910). The conceptual domains of the PSMA are consistent with internationally recognized nursing safety activity frameworks, including the World Health Organization patient safety curriculum and evidence-based hospital safety practices [23], [24].

#### Patient safety culture

PSC was assessed using the Hospital Survey on PSC (HSOPSC), developed by the Agency for Healthcare Research and Quality (AHRQ). The HSOPSC consists of 42 items across 12 dimensions and is rated on a 5-point Likert scale. Eighteen items are reverse-coded. Positive response rates were calculated following AHRQ guidelines, and mean scores were used for regression analyses. The Vietnamese version of the HSOPSC was translated by Ho Chi Minh city Department of Health and has been officially authorized for public use in Vietnam. This version has demonstrated acceptable reliability and validity in hospital settings. In this study, the overall Cronbach’s alpha for the HSOPSC was 0.889 [25].

### Statistical analysis

Data were analyzed using SPSS software version 20.0 (IBM, Armonk, NY, USA). Descriptive statistics were used to summarize participant characteristics and study variables. Internal consistency reliability was assessed using Cronbach’s alpha coefficients. Pearson correlation analysis was conducted to examine bivariate associations among study variables. Multivariable linear regression analyses were performed to test the hypothesized conceptual framework. In Model 1, PCC competence (P-CAT) and PSCP (H-PEPSS) were entered as predictors of PSMA. In Model 2, PCC competence and PSCp were entered as predictors of patient safety culture (HSOPSC). In Model 3, PSMA was added to Model 2 to examine its mediating role. Sociodemographic and professional variables (age, gender, clinical experience, education level, training in patient safety, job satisfaction, and weekly working hours) were included as covariates in all models. Standardized beta coefficients (β), p-values, R², and F statistics were reported [26].

Mediation analyses were conducted using bootstrapping with 5,000 resamples to estimate indirect effects and 95% bias-corrected confidence intervals. Indirect effects were considered statistically significant if the confidence interval did not include zero [27]. Statistical significance was set at p < 0.05 (two-tailed).

### Ethical considerations

The study was approved by the Ethics Committee of Hue University of Medicine and Pharmacy (No. H2024/022). Permission was obtained from participating hospitals prior to data collection. Participation was voluntary and anonymous. Written informed consent was obtained from all participants before enrollment. All data were collected and analyzed in a manner that ensured confidentiality and anonymity throughout the study.

## Results

A total of 1,036 nurses were included in the analysis, yielding a response rate of 99.6%. The mean age of participants was 35.41 ± 6.45 years (range: 22–56), and the majority were older than 30 years (77.5%). Most participants were female (89.8%), of Kinh ethnicity (97.3%), and reported no religious affiliation (92.9%). Approximately half of the nurses held a diploma or associate degree (50.9%), while 49.1% had a bachelor’s degree or higher. Most participants were married (82.8%) and worked as staff nurses (92.8%). The mean duration of clinical experience was 12.26 ± 6.31 years, and the mean number of years working in the current department was 10.28 ± 6.00 years. The average weekly working time was 52.67 ± 13.89 hours, with 62.2% working more than 40 hours per week. Nurses worked a mean of 6.26 ± 3.29 night shifts per month. Most participants reported being satisfied with their work (80.1%). Nearly all nurses had experience with patient safety education (99.9%) and PCC education (99.2%). The majority reported experience in patient safety practice (99.5%) and PCC practice (98.0%). A high proportion had experienced patient safety incidents (91.6%), and 66.0% had reported such incidents. Most participants indicated that incident reporting was encouraged in their workplace (98.2%). Overall, 82.7% rated PSC as good, and 74.8% perceived patient safety in their department as good.

**Table 1.** General and work-related characteristics of the participants (N=1036)

| Characteristics |  | Frequency (n) | Percentage (%) |
| --- | --- | --- | --- |
| | Mean ± SD = 35,41 ± 6,45 (22-56) | | |
| **Age** | $\leq 30$ | 233 | 22.50 |
| | $> 30$ | 803 | 77.50 |
| **Gender** | Male | 106 | 10.20 |
|  | Female | 930 | 89.80 |
| **Ethnicity** | Kinh | 1008 | 97.30 |
|  | Other | 28 | 2.70 |
| **Religion** | No | 962 | 92.90 |
|  | Yes | 74 | 7.10 |
| **Education level** | Diploma/Associate degree | 527 | 50.90 |
|  | Bachelor's degree or higher | 509 | 49.10 |
| **Marital status** | Single | 151 | 14.60 |
|  | Married | 858 | 82.80 |
|  | Other | 27 | 2.60 |
| **Position** | Staff | 961 | 92.80 |
|  | Head nurse | 75 | 7.20 |
| **Years of clinical experience (year)** | Mean ± SD = 12.26 ± 6.31 (1-35) | | |
|  | < 5 | 69 | 6.70 |
|  | 5–9 | 327 | 31.60 |
|  | 10–14 | 305 | 29.40 |
| | ≥ 15 | 335 | 32.30 |
| **Years working at current department (year)** | Mean ± SD = 10.28 ± 6.00 (0.3-35) | | |
|  | < 5 | 185 | 17.90 |
|  | 5–9 | 330 | 31.90 |
|  | 10–14 | 270 | 26.10 |
|  | ≥ 15 | 251 | 24.20 |
| **Weekly working hours (hours)** | Mean ± SD = 52.67 ± 13.89 (40-102) |  |  |
|  | ≤ 40 | 392 | 37.80 |
|  | > 40 | 644 | 62.20 |
| **Night shifts per month** | Mean ± SD = 6.26 ± 3.29 (0-12) |  |  |
|  | < 5 | 243 | 23.50 |
|  | 5–9 | 624 | 60.20 |
|  | ≥ 10 | 169 | 16.30 |
| **Satisfied with work** | Dissatisfied | 206 | 19.90 |
|  | Satisfied | 830 | 80.10 |
| **Experience of patient safety education** | Yes | 1035 | 99.90 |
|  | No | 1 | 0.10 |
| **Experience of PCC education** | Yes | 1028 | 99.20 |
|  | No | 8 | 0.80 |
| **Experience of patient safety practice** | Yes | 1031 | 99.50 |
|  | No | 5 | 0.50 |
| **Experience of PCC practice** | Yes | 1015 | 98.00 |
|  | No | 21 | 2.00 |
| **Patient safety incident experience** | Yes | 949 | 91.60 |
|  | No | 87 | 8.40 |
| **Patient safety incident report** | Yes | 684 | 66.00 |
|  | No | 352 | 34.00 |
| **Encouragement for incident reporting** | Yes | 1017 | 98.20 |
|  | No | 19 | 1.80 |
| **Self-assessment of PSC** | Not good | 179 | 17.30 |
|  | Good | 857 | 82.7 |
| **Self-rated patient safety in the department** | Not good | 261 | 25.20 |
|  | Good | 775 | 74.80 |

Differences in person-centered care competence, PSCp, PSMA, and PSC across general and work-related characteristics are presented in **Table 2**. Significant variations were observed across multiple demographic and professional factors (all p < 0.05). Higher PCC competence was reported by female nurses, head nurses, those working ≤40 hours per week, nurses satisfied with their work, those with PCC education and practice experience, and those reporting favorable safety conditions. PSCp varied significantly by age, education, department, position, clinical experience, night shift frequency, job satisfaction, safety-related experiences, and self-rated safety indicators, with generally higher scores among older, more experienced, and more highly educated nurses in leadership roles.

**Table 2.** Differences in person-centered care, patient safety competence, patient safety nursing activities, and patient safety culture according to general and work-related characteristics (N=1036)

| Variables | Categories | PCAT |  | HPEPSS |  | PSMA |  | HSOPSC |  |
| --- | --- | --- | --- | --- | --- | --- | --- | --- | --- |
| | | Mean ±<br>SD | p<br>*Scheffe's*<br>*test* | Mean ±<br>SD | p<br>*Scheffe's*<br>*test* | Mean ±<br>SD | p<br>*Scheffe's*<br>*test* | Mean ±<br>SD | p<br>*Scheffe's*<br>*test* |
| Age | $\leq 30$ | 3.78±0.36 | 0.617 | 4.17±0.36 | **0.004** | 4.44±0.35 | 0.839 | 3.93±0.38 | 0.767 |
| | | Mean ±<br>SD | p<br>*Scheffe's*<br>*test* | Mean ±<br>SD | p<br>*Scheffe's*<br>*test* | Mean ±<br>SD | p<br>*Scheffe's*<br>*test* | Mean ±<br>SD | p<br>*Scheffe's*<br>*test* |
| | > 30 | 3.77±0.36 | | 4.24±0.36 | | 4.44±0.35 | | 3.94±0.34 | |
| Gender | Male | 3.66±0.33 | **0.001** | 4.25±0.36 | 0.690 | 4.39±0.36 | 0.161 | 3.82±0.38 | **0.001** |
| | Female | 3.78±0.37 | | 4.23±0.36 | | 4.44±0.35 | | 3.95±0.35 | |
| Ethnicity | Kinh | 3.77±0.36 | 0.940 | 4.23±0.37 | 0.061 | 4.44±0.35 | **0.005** | 3.93±0.35 | 0.862 |
| | Other | 3.77±0.29 | | 4.14±0.26 | | 4.25±0.37 | | 3.92±0.21 | |
| Religion | No | 3.78±0.37 | 0.130 | 4.23±0.36 | 0.992 | 4.44±0.35 | 0.068 | 3.94±0.35 | 0.111 |
| | Yes | 3.71±0.32 | | 4.23±0.37 | | 4.36±0.37 | | 3.87±0.35 | |
| Education level | Diploma/Associate degree | 3.77±0.37 | 0.940 | 4.20±0.37 | **0.003** | 4.43±0.35 | 0.285 | 3.92±0.35 | 0.220 |
|  |  | Mean ± SD | p<br>*Scheffe's test* | Mean ± SD | p<br>*Scheffe's test* | Mean ± SD | p<br>*Scheffe's test* | Mean ± SD | p<br>*Scheffe's test* |
|  | Bachelor's degree or higher | 3.77±0.36 |  | 4.27±0.36 |  | 4.45±0.36 |  | 3.95±0.35 |  |
| Marital status | Single | 3.75±0.37 | 0.618 | 4.17±0.39 | **0.047** | 4.44±0.35 | 0.587 | 3.87±0.41 | **0.046** |
|  | Married | 3.77±0.36 |  | 4.24±0.36 |  | 4.43±0.36 |  | 3.94±0.34 |  |
|  | Other | 3.82±0.37 |  | 4.28±0.36 |  | 4.50±0.29 |  | 3.98±0.45 |  |
| Working department | Internal medicine<sup>a</sup> | 3.77±0.35 | <**0.001**<br>a,b<e | 4.30±0.35 | <**0.001**<br>b,c<a<br>b<e | 4.47±0.37 | <**0.001**<br>b<a,d | 4.02±0.35 | <**0.001**<br>b,d<a<br>b<c<br>b<e |
|  | Surgery<sup>b</sup> | 3.74±0.35 |  | 4.16±0.35 |  | 4.34±0.34 |  | 3.82±0.32 |  |
|  | Obstetrics–<br>Pediatrics–<br>Oncology<sup>c</sup> | 3.80±0.38 |  | 4.16±0.40 |  | 4.41±0.37 |  | 3.94±0.32 |  |
|  |  | Mean ± SD | p<br>*Scheffe's test* | Mean ± SD | p<br>*Scheffe's test* | Mean ± SD | p<br>*Scheffe's test* | Mean ± SD | p<br>*Scheffe's test* |
|  | Anesthesiology-ICU-Emergency | 3.71±0.37 |  | 4.23±0.37 |  | 4.50±0.32 |  | 3.86±0.39 |  |
|  | Other<sup>e</sup> | 3.91±0.36 |  | 4.29±0.33 |  | 4.45±0.35 |  | 4.06±0.28 |  |
| Position | Staff | 3.76±0.36 | **0.002** | 4.22±0.37 | **0.001** | 4.44±0.35 | 0.854 | 3.93±0.35 | 0.278 |
|  | Head nurse | 3.90±0.33 |  | 4.36±0.33 |  | 4.44±0.35 |  | 3.98±0.32 |  |
| Years of clinical experience (year) | < 5<sup>a</sup> | 3.79±0.33 | 0.759 | 4.13±0.32 | <**0.001**<br>a,b<d | 4.43±0.32 | 0.538 | 3.94±0.38 | 0.208 |
|  | 5-<10<sup>b</sup> | 3.77±0.38 |  | 4.20±0.37 |  | 4.43±0.36 |  | 3.91±0.37 |  |
|  | 10-<15<sup>c</sup> | 3.75±0.35 |  | 4.22±0.37 |  | 4.42±0.36 |  | 3.92±0.34 |  |
|  | ≥ 15<sup>d</sup> | 3.78±0.37 |  | 4.30±0.35 |  | 4.46±0.34 |  | 3.97±0.34 |  |
|  | < 5<sup>a</sup> | 3.86±0.34 | **0.003** | 4.21±0.34 | 0.063 | 4.40±0.35 | 0.273 | 3.97±0.33 | 0.197 |
|  |  | Mean ± SD | p<br>*Scheffe's test* | Mean ± SD | p<br>*Scheffe's test* | Mean ± SD | p<br>*Scheffe's test* | Mean ± SD | p<br>*Scheffe's test* |
| Years working at current department (year) | 5–<10<sup>b</sup> | 3.77±0.38 | c,d<a | 4.21±0.37 |  | 4.43±0.36 |  | 3.91±0.37 |  |
|  | 10–<15<sup>c</sup> | 3.74±0.34 |  | 4.23±0.38 |  | 4.47±0.35 |  | 3.91±0.34 |  |
|  | ≥ 15<sup>d</sup> | 3.75±0.37 |  | 4.28±0.35 |  | 4.44±0.34 |  | 3.95±0.35 |  |
| Weekly working hours (hours) | ≤ 40 | 3.80±0.36 | **0.024** | 4.24±0.36 | 0.418 | 4.41±0.36 | 0.084 | 3.98±0.33 | **0.002** |
|  | > 40 | 3.75±0.36 |  | 4.22±0.37 |  | 4.45±0.35 |  | 3.91±0.36 |  |
| Night shifts per month | < 5<sup>a</sup> | 3.81±0.36 | 0.074 | 4.28±0.34 | **0.012**<br>c<a | 4.41±0.37 | 0.240 | 3.99±0.34 | <**0.001**<br>c<a,b |
|  | 5–<10<sup>b</sup> | 3.77±0.36 |  | 4.23±0.37 |  | 4.45±0.35 |  | 3.95±0.35 |  |
|  | ≥ 10<sup>c</sup> | 3.73±0.38 |  | 4.17±0.35 |  | 4.41±0.33 |  | 3.77±0.33 |  |
| Satisfied with work | Dissatisfied | 3.62±0.35 | <**0.001** | 4.10±0.38 | <**0.001** | 4.39±0.38 | **0.045** | 3.78±0.39 | <**0.001** |
|  | Satisfied | 3.81±0.36 |  | 4.27±0.35 |  | 4.45±0.35 |  | 3.97±0.33 |  |
|  | Yes | 3.77±0.36 | **0.028** | 4.23±0.36 | **0.023** | 4.44±0.35 | 0.765 | 3.39±0.35 | 0.146 |
|  | No | 3.49±0.30 |  | 3.94±0.19 |  | 4.47±0.28 |  | 3.75±0.31 |  |
|  |  | Mean ± SD | p<br>*Scheffe's test* | Mean ± SD | p<br>*Scheffe's test* | Mean ± SD | p<br>*Scheffe's test* | Mean ± SD | p<br>*Scheffe's test* |
| Experience of PCC education |  |  |  |  |  |  |  |  |  |
| Experience of patient safety practice | Yes | 3.77±0.36 | 0.580 | 4.23±0.36 | 0.159 | 4.44±0.35 | **0.034** | 3.93±0.35 | 0.951 |
|  | No | 3.86±0.30 |  | 4.46±0.28 |  | 4.58±0.11 |  | 3.94±0.20 |  |
| Experience of PCC practice | Yes | 3.78±0.36 | **0.023** | 4.24±0.36 | <**0.001** | 4.44±0.35 | 0.715 | 3.94±0.35 | **0.021** |
|  | No | 3.59±0.29 |  | 4.01±0.24 |  | 4.41±0.35 |  | 3.76±0.26 |  |
| Patient safety incident experience | Yes | 3.77±0.37 | 0.512 | 4.24±0.37 | <**0.001** | 4.44±0.35 | 0.871 | 3.94±0.36 | <**0.001** |
|  | No | 3.75±0.33 |  | 4.09±0.32 |  | 4.44±0.34 |  | 3.81±0.27 |  |
|  | Yes | 3.78±0.35 | 0.280 | 4.26±0.35 | **0.003** | 4.45±0.33 | 0.303 | 3.96±0.34 | <**0.001** |
|  |  | Mean ± SD | p<br>*Scheffe's test* | Mean ± SD | p<br>*Scheffe's test* | Mean ± SD | p<br>*Scheffe's test* | Mean ± SD | p<br>*Scheffe's test* |
| Patient safety incident report | No | 3.75±0.39 |  | 4.19±0.38 |  | 4.42±0.39 |  | 3.87±0.37 |  |
| Encouragement for incident reporting | Yes | 3.77±0.37 | 0.177 | 4.23±0.36 | 0.501 | 4.44±0.35 | 0.298 | 3.94±0.35 | 0.083 |
|  | No | 3.67±0.26 |  | 4.18±0.40 |  | 4.35±0.32 |  | 3.79±0.38 |  |
| Self-assessment of PSC | Not good | 3.63±0.32 | <0.001 | 4.08±0.35 | <0.001 | 4.29±0.37 | <0.001 | 3.75±0.32 | <0.001 |
|  | Good | 3.80±0.37 |  | 4.26±0.36 |  | 4.47±0.34 |  | 3.97±0.35 |  |
| Self-rated patient safety in the department | Not good | 3.70±0.35 | <0.001 | 4.10±0.31 | <0.001 | 4.37±0.35 | <0.001 | 3.76±0.30 | <0.001 |
|  | Good | 3.80±0.37 |  | 4.28±0.37 |  | 4.46±0.35 |  | 3.99±0.35 |  |

PSMA and PSC also differed significantly across several work-related factors, particularly department, night shifts, job satisfaction, and safety perceptions, with consistently higher scores among nurses reporting more favorable work environments and safety climates.

Descriptive statistics and correlations among PCC competence PSC, PSMA, and PSC are presented in **Table 3**. The mean scores were 3.77 ± 0.36 for P-CAT, 4.23 ± 0.36 for H-PEPSS, 4.44 ± 0.35 for PSMA, and 3.93 ± 0.35 for HSOPSC. Pearson correlation analysis indicated that P-CAT was positively correlated with H-PEPSS (r = 0.32, p < 0.001), PSMA (r = 0.24, p < 0.001), and HSOPSC (r = 0.49, p < 0.001). H-PEPSS was also positively associated with PSMA (r = 0.35, p < 0.001) and HSOPSC (r = 0.33, p < 0.001). PSMA showed a significant positive correlation with HSOPSC (r = 0.27, p < 0.001). All correlation coefficients were in the low-to-moderate range.

**Table 3.** Descriptive analysis and correlation of person-centered care, patient safety competence, patient safety nursing activities, and patient safety culture (N=1036)

| Variable | Mean ± SD | Min - Max | PCAT | HPEPSS | PSMA | HSOPSC |
| --- | --- | --- | --- | --- | --- | --- |
|  |  |  | r<br>(p-value) | r<br>(p-value) | r<br>(p-value) | r<br>(p-value) |
| PCAT | 3.77 ± 0.36 | 2.77 - 4.85 | 1 |  |  |  |
| HPEPSS | 4.23 ± 0.36 | 3.09 - 5.00 | 0.32<br>($< 0.001$) | 1 | | |
| PSMA | 4.44 ± 0.35 | 3.25 - 5.00 | 0.24<br>($< 0.001$) | 0.35<br>($< 0.001$) | 1 | |
| HSOPSC | 3.93 ± 0.35 | 2.81 - 4.79 | 0.49<br>($< 0.001$) | 0.33<br>($< 0.001$) | 0.27<br>($< 0.001$) | 1 |

*Notes: Pearson’s correlation analysis*

Multivariable regression analyses examining predictors of PSMA and PSC (HSOPSC) are presented in **Table 4**. In Model 1, both PCC competence (P-CAT; β = 0.149, p < 0.001) and PSC (H-PEPSS; β = 0.274, p < 0.001) were independently associated with PSMA after adjustment for covariates, explaining 18.2% of the variance (adjusted R² = 0.173). In Model 2, P-CAT (β = 0.390, p < 0.001) and H-PEPSS (β = 0.108, p < 0.001) were significant predictors of HSOPSC, with the model accounting for 37.7% of the variance (adjusted R² = 0.366). Several work-related factors, including working department, frequent night shifts (≥10 per month), patient safety incident experience, self-assessment of PSC, and self-rated patient safety in the department, were also significantly associated with HSOPSC. In Model 3, when PSMA was included, PSMA was independently associated with HSOPSC (β = 0.102, p < 0.001), while the regression coefficients for both P-CAT (β = 0.375, p < 0.001) and H-PEPSS (β = 0.079, p = 0.006) were attenuated but remained statistically significant, indicating partial mediation. The final model explained 38.6% of the variance in PSC (adjusted R² = 0.374).

**Table 4.**
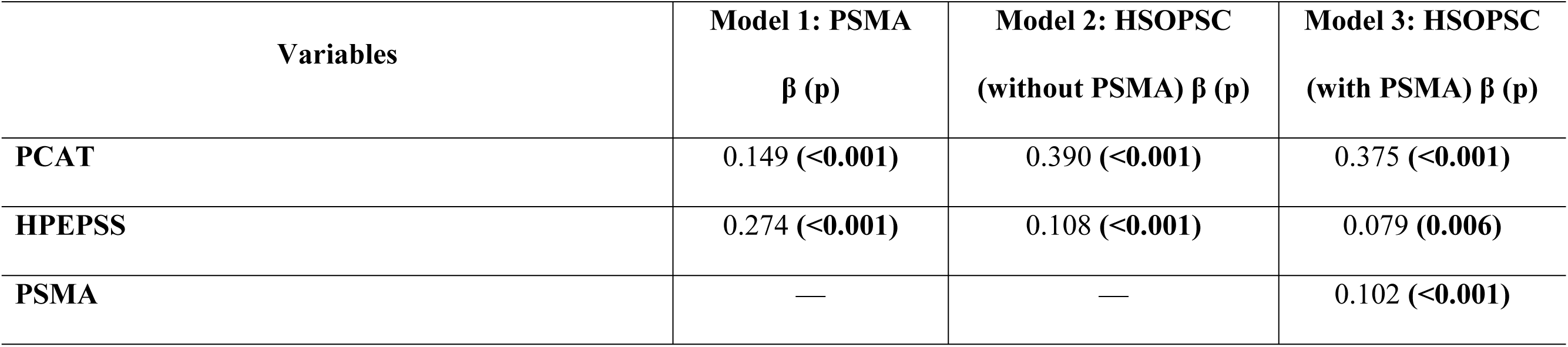
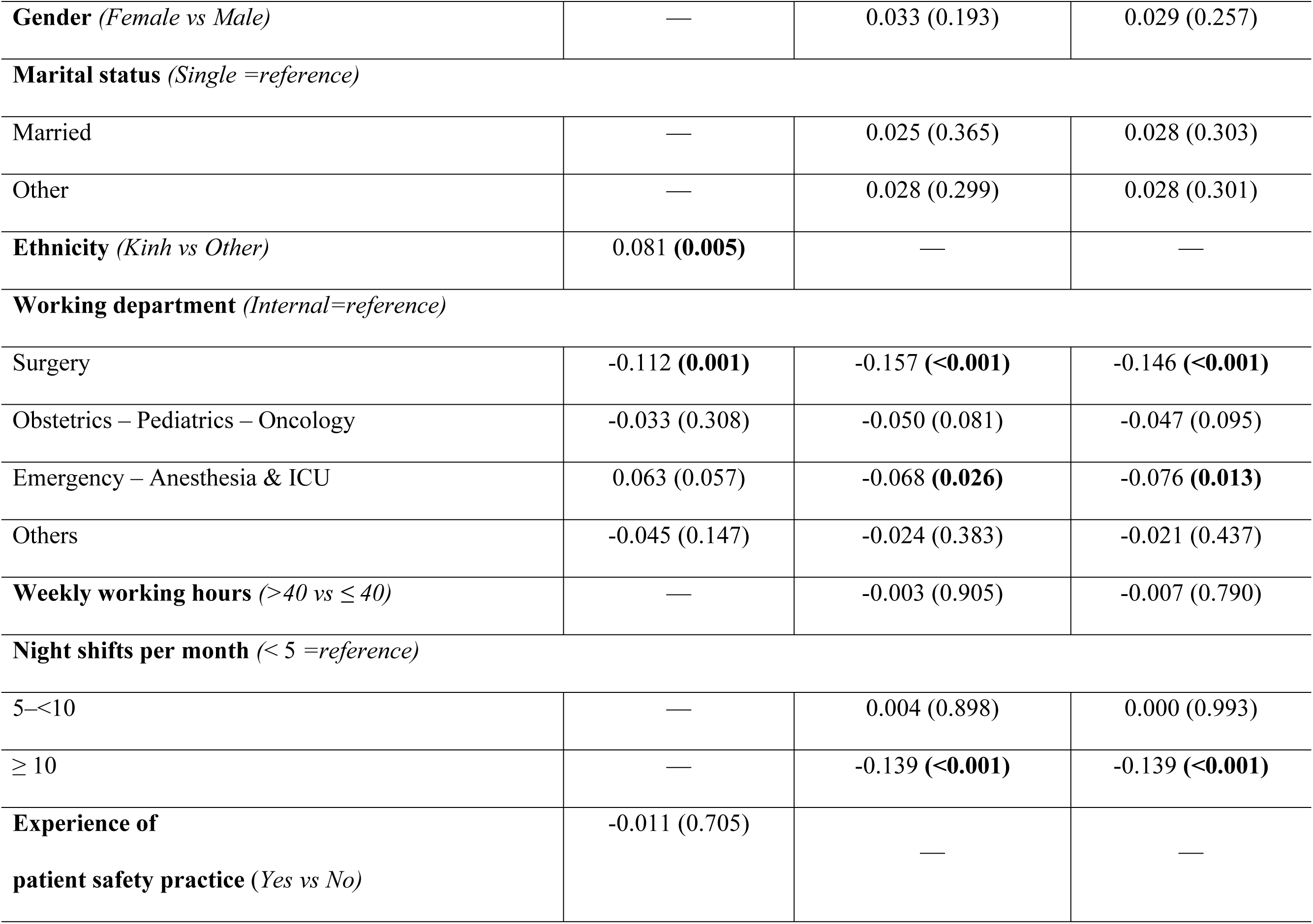
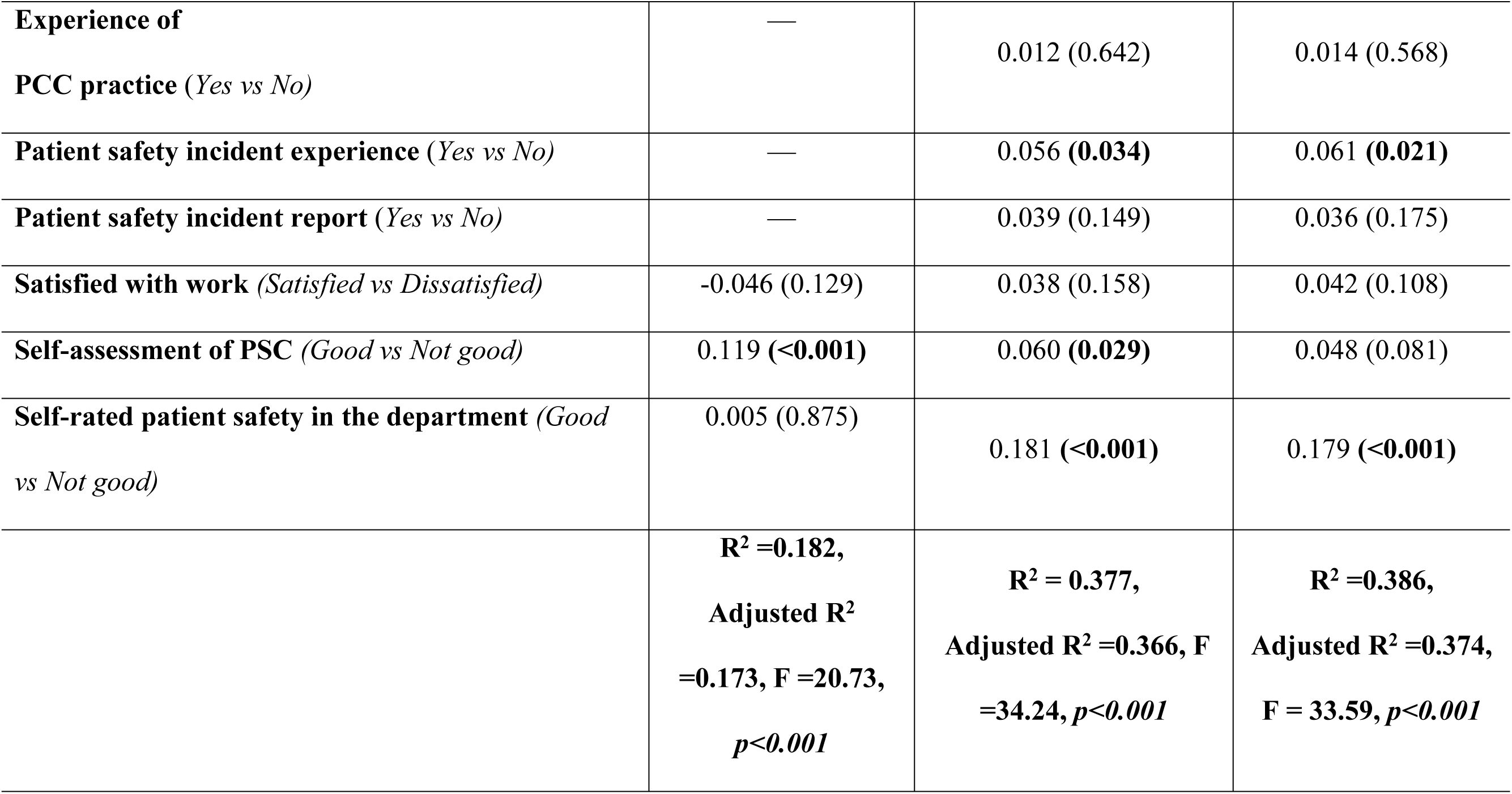
Multivariable regression models predicting patient safety competence, patient safety nursing activities, and patient safety culture (N=1036)

| Variables | Model 1: PSMA<br>$\beta$ (p) | Model 2: HSOPSC<br>(without PSMA) $\beta$ (p) | Model 3: HSOPSC<br>(with PSMA) $\beta$ (p) |
| --- | --- | --- | --- |
| PCAT | 0.149 (<0.001) | 0.390 (<0.001) | 0.375 (<0.001) |
| HPEPSS | 0.274 (<0.001) | 0.108 (<0.001) | 0.079 (0.006) |
| PSMA | — | — | 0.102 (<0.001) |
| **Gender** (*Female vs Male*) | — | 0.033 (0.193) | 0.029 (0.257) |
| **Marital status** (*Single =reference*) |  |  |  |
| Married | — | 0.025 (0.365) | 0.028 (0.303) |
| Other | — | 0.028 (0.299) | 0.028 (0.301) |
| **Ethnicity** (*Kinh vs Other*) | 0.081 (**0.005**) | — | — |
| **Working department** (*Internal=reference*) |  |  |  |
| Surgery | -0.112 (**0.001**) | -0.157 (<**0.001**) | -0.146 (<**0.001**) |
| Obstetrics – Pediatrics – Oncology | -0.033 (0.308) | -0.050 (0.081) | -0.047 (0.095) |
| Emergency – Anesthesia & ICU | 0.063 (0.057) | -0.068 (**0.026**) | -0.076 (**0.013**) |
| Others | -0.045 (0.147) | -0.024 (0.383) | -0.021 (0.437) |
| **Weekly working hours** (*>40 vs ≤ 40*) | — | -0.003 (0.905) | -0.007 (0.790) |
| **Night shifts per month** (*< 5 =reference*) |  |  |  |
| 5–<10 | — | 0.004 (0.898) | 0.000 (0.993) |
| ≥ 10 | — | -0.139 (<**0.001**) | -0.139 (<**0.001**) |
| **Experience of patient safety practice** (*Yes vs No*) | -0.011 (0.705) | — | — |
| **Experience of PCC practice** (*Yes vs No*) | — | 0.012 (0.642) | 0.014 (0.568) |
| **Patient safety incident experience** (*Yes vs No*) | — | 0.056 (**0.034**) | 0.061 (**0.021**) |
| **Patient safety incident report** (*Yes vs No*) | — | 0.039 (0.149) | 0.036 (0.175) |
| **Satisfied with work** (*Satisfied vs Dissatisfied*) | -0.046 (0.129) | 0.038 (0.158) | 0.042 (0.108) |
| **Self-assessment of PSC** (*Good vs Not good*) | 0.119 (**<0.001**) | 0.060 (**0.029**) | 0.048 (0.081) |
| **Self-rated patient safety in the department** (*Good vs Not good*) | 0.005 (0.875) | 0.181 (**<0.001**) | 0.179 (**<0.001**) |
|  | **R<sup>2</sup> =0.182,**<br>**Adjusted R<sup>2</sup>**<br>**=0.173, F =20.73,**<br>***p*<0.001** | **R<sup>2</sup> = 0.377,**<br>**Adjusted R<sup>2</sup> =0.366, F**<br>**=34.24, *p*<0.001** | **R<sup>2</sup> =0.386,**<br>**Adjusted R<sup>2</sup> =0.374,**<br>**F = 33.59, *p*<0.001** |

Mediation analysis using bootstrapping with 5,000 resamples demonstrated that PSMA partially mediated the relationships between both PCC competence and PSC, as well as PSCp and PSC (Table 5). For the pathway from PCC competence (P-CAT) to PSC (HSOPSC), the total effect was statistically significant (β = 0.4055, 95% CI: 0.3561–0.4549). After accounting for PSMA, the direct effect remained significant (β = 0.3790, 95% CI: 0.3288–0.4291), while the indirect effect through PSMA was also significant (β = 0.0265, 95% CI: 0.0143–0.0404), indicating partial mediation. Similarly, PSCp (H-PEPSS) showed a significant total effect on PSC (β = 0.2125, 95% CI: 0.1577–0.2673). When PSMA was included in the model, the direct effect was attenuated but remained significant (β = 0.1628, 95% CI: 0.1057–0.2199), and a significant indirect effect via PSMA was observed (β = 0.0497, 95% CI: 0.0304–0.0711), also supporting partial mediation. In both models, the indirect effects were statistically significant as the bias-corrected confidence intervals did not include zero, suggesting that PSMA represent an important behavioral pathway through which nurses’ competencies are associated with PSC.

**Table 5.** Mediation effects of patient safety management activities in the relationships between professional competencies and patient safety culture (Bootstrapping with 5,000 resamples) (N=1036)

| Pathway | Effect type | $\beta$ | p-value | 95% CI |
| --- | --- | --- | --- | --- |
| P-CAT → PSMA → HSOPSC | Total effect (c) | 0.4055 | <0.001 | 0.3561 – 0.4549 |
|  | Direct effect (c') | 0.3790 | <0.001 | 0.3288 – 0.4291 |
|  | Indirect effect (a×b) | 0.0265 | — | 0.0143 – 0.0404 |
| **H-PEPSS → PSMA →**<br>**HSOPSC** | Total effect (c) | 0.2125 | <0.001 | 0.1577 – 0.2673 |
|  | Direct effect (c') | 0.1628 | <0.001 | 0.1057 – 0.2199 |
|  | Indirect effect (a×b) | 0.0497 | — | 0.0304 – 0.0711 |

*Note: Indirect effects were estimated using bias-corrected bootstrapping with 5,000 resamples. Indirect effects were considered statistically significant when the 95% confidence interval did not include zero. p-values are not reported for indirect effects, in accordance with recommendations for bootstrap mediation analysis.*

## DISCUSSION

### Levels of PCC competence, patient safety competence, patient safety management activities, and patient safety culture

The present findings indicate that nurses reported moderate-to-high levels of PCC competence, PSCp, and PSMA, whereas PSC was perceived at a moderate level. This pattern—relatively strong individual competence alongside only moderate organizational safety culture—has been observed in several hospital settings, particularly in low- and middle-income countries [28], [29], [30].

This divergence may reflect the distinction between individual capability and collective organizational climate. While professional competence is largely shaped by education, training, and clinical exposure, safety culture is embedded within institutional structures, leadership behaviors, communication norms, and reporting systems. Evidence from validation studies of PSCp instruments suggests that nurses often rate interpersonal domains, such as teamwork and communication, more positively than system-level dimensions related to organizational safety infrastructure [9], [31].

The relatively high level of engagement in PSMA observed in this study further supports the interpretation that nurses consistently perform protocol-driven safety behaviors embedded in daily workflows. Similar findings have been reported in studies using validated safety activity scales [22], In Vietnamese hospital settings, previous studies have also reported generally positive engagement with patient safety practices and safety culture among nurses [32].

However, despite these favorable behavioral indicators, PSC remained moderate. This gap has been consistently described in the safety literature, where healthcare professionals frequently report adequate knowledge and compliance with safety practices while expressing less confidence in institutional safety systems [2]. Vietnamese hospital studies have similarly highlighted variability across culture domains, particularly in staffing adequacy, non-punitive response to error, and open communication [32].

From a conceptual perspective, these findings align with a structure–process–outcome model, in which professional competence represents a structural input, patient safety management activities reflect care processes, and PSC constitutes an organizational outcome [14]. Global patient safety policy frameworks emphasize that improvements in competence and safety practices require supportive leadership, learning systems, and reporting environments to generate sustained cultural change [4], [5].

These findings underscore a structural tension between individual professional preparedness and organizational readiness for patient safety within tertiary hospital settings.

### Direct and indirect relationships among competencies, safety activities, and patient safety culture

Beyond descriptive findings, this study examined how PCC competence and PSCp are linked to PSC through both direct and indirect pathways. Both competence domains were significantly associated with PSC, and PSMA partially mediated these relationships. The significant indirect effects observed in the bootstrapped mediation analyses indicate that frontline safety behaviors represent an important mechanism through which professional competencies are translated into broader organizational safety culture.

The direct association between PSCp and PSC observed in this study is consistent with accumulating international evidence demonstrating that higher levels of clinical competence are associated with stronger safety culture across hospital settings [11]. Multicenter cross-sectional research has further shown that PSCp is positively associated with safety culture and inversely associated with adverse events, suggesting that competence contributes not only to perceptions but also to measurable safety outcomes [33]. These findings reinforce the interpretation that competence in teamwork, communication, and risk management contributes to shaping shared safety norms within clinical environments. Importantly, the mediating role of PSMA supports a competence–behavior–culture pathway. PSMA represent observable and routine safety practices embedded in daily nursing workflows, including patient identification, medication safety, infection prevention, and fall prevention. Evidence indicates that safety competence and safety culture jointly predict safety nursing activities [34], and that safety culture may mediate relationships between professional attributes and safety-related outcomes [35]. The present findings extend this body of work by demonstrating that frontline safety behaviors function as a practical conduit through which competence is operationalized and subsequently reflected in collective safety perceptions.

The partial nature of the mediation effect is theoretically meaningful. While engagement in safety management activities explains part of the association between competence and PSC, a substantial direct effect remained. This suggests that competencies influence safety culture not only through behavioral enactment but also through cognitive and attitudinal mechanisms, such as risk awareness, professional responsibility, and shared commitment to safety. At the same time, extensive evidence indicates that safety culture is shaped by broader organizational determinants, including leadership engagement, communication climate, staffing adequacy, and non-punitive responses to error [36], [30].

Within the Vietnamese context, national studies have similarly demonstrated that managerial support, communication openness, and reporting systems significantly influence safety culture perceptions [32], [28]. These contextual findings support the interpretation that individual competence, although necessary, is insufficient in isolation without supportive organizational conditions that enable consistent and psychologically safe enactment of safety practices.

Taken together, the present findings support an integrated and multidimensional pathway linking nurses’ competencies, frontline safety behaviors, and patient safety culture. Strengthening PCC competence and PSCp may enhance safety culture both directly and indirectly through sustained engagement in PSMA. However, durable cultural transformation likely requires simultaneous reinforcement of leadership practices, organizational learning systems, and supportive work environments to align individual capability with institutional readiness for patient safety.

## Conclusion

This study demonstrates that PCC competence and PSCp are significantly associated with PSC among nurses in tertiary hospitals. PSMA partially mediated these relationships, indicating that frontline safety behaviors play an important role in translating individual competencies into organizational safety culture.

While strengthening professional competencies may contribute to improving PSC, sustainable cultural change also requires supportive organizational conditions, including effective leadership and enabling work environments. Efforts to enhance PSC should therefore integrate competence development with system-level reinforcement of safety practices.

Future longitudinal and interventional studies are warranted to clarify causal pathways and test system-level strategies for strengthening PSC.

## Data Availability

All relevant data are within the manuscript and its Supporting Information files

## Author contributions

Conceptualization: Duong Thi Ngoc Lan, Ho Duy Binh.

Data curation: Dang Thi Thanh Phuc, Vo Thi Nhi, Ho Thi My Yen, Nguyen Binh Thao Nguyen, Tran Thi Kim Quyen.

Formal analysis: Dang Thi Thanh Phuc.

Investigation: Dang Thi Thanh Phuc, Vo Thi Nhi, Ho Thi My Yen, Nguyen Binh Thao Nguyen, Tran Thi Kim Quyen.

Methodology: Duong Thi Ngoc Lan, Ho Duy Binh, Dang Thi Thanh Phuc, Vo Thi Nhi, Ho Thi My Yen.

Supervision: Duong Thi Ngoc Lan, Ho Duy Binh.

Writing – original draft: Duong Thi Ngoc Lan, Ho Duy Binh, Dang Thi Thanh Phuc, Ho Thi My Yen.

Writing – review & editing: Duong Thi Ngoc Lan, Ho Duy Binh.

All authors have read and agreed to the published version of the manuscript.

## Supporting Information

S1 Dataset. Mediating effects of patient safety management activities among nurses dataset. (.sav)

## Acknowledgments

The authors would like to thank the participating hospitals and all nurses who contributed to this study. We also acknowledge the support of the nursing departments at the participating institutions for facilitating data collection.

## Funding

This research was funded by the Ministry of Education and Training of Vietnam under grant number: B2024-DHH-04. The funder had no role in study design, data collection and analysis, decision to publish, or preparation of the manuscript.

## Notes

### Competing Interest Statement

The authors have declared no competing interest.

### Funding Statement

Yes

### Author Declarations

Approval number: H2024/022 February 5th, 2024 Ms. Ho Duy Binh Hue University of Medicine and Pharmacy, Hue University, Vietnam Subject: Approval of the study: “Patient safety competence and patient safety culture among nurses working in level I hospitals in Central of Vietnam”. Dear Mr. Ho Duy Binh, The Institutional Ethics Committee of Hue University of Medicine and Pharmacy has reviewed and approved the following study: Study title: Patient safety competence and patient safety culture among nurses working in level I hospitals in Central of Vietnam Principle Investigator: Nguyen Ngoc Quynh Anh, Office of Student Affairs, Hue University of Medicine and Pharmacy, Hue University, Vietnam. Co-investigators: Duong Thi Ngoc Lan, Faculty of Nursing, Hue University of Medicine and Pharmacy, Hue University, Vietnam. Dang Thi Thanh Phuc, Faculty of Nursing, Hue University of Medicine and Pharmacy, Hue University, Vietnam. Vo Thi Nhi, Faculty of Nursing, Hue University of Medicine and Pharmacy, Hue University, Vietnam. This project is approved for the research period from February 2024 to December 2025. It is your responsibility to ensure that all people associated with the study are made aware of what has been actually approved. HUE UNIVERSITY OF MEDICINE AND PHARMACY 06 Ngo Quyen street, Hue city – VIET NAM Telephone: +84.234.3822873 Fax: +84.234.3826269 Website: huemed-univ.edu.vnPlease note that the following conditions apply to your approval. Failure to abide by these conditions may result in suspension or discontinuation of approval and/or disciplinary action. a. Limit of Approval: Approval is limited strictly to the study as submitted in your application b. All procedures within this study must follow what have been submitted in your ethics application. c. Approval is for the above mentioned period. Research must be renewed (if needed) until it is complete. Yours sincerely, The Institutional Ethics Committee of Hue University of Medicine and Pharmacy CHAIR Prof. VO TAM SECRETARY A/Prof. NGUYEN THANH THAO

## References

1. Weaver SJ, Lubomksi LH, Wilson RF, Pfoh ER, Martinez KA, Dy SM. Promoting a culture of safety as a patient safety strategy: a systematic review. Ann Intern Med. 2013;158(5 Pt 2):369-74. doi: 10.7326/0003-4819-158-5-201303051-00002.

2. Halligan M, Zecevic A. Safety culture in healthcare: a review of concepts, dimensions, measures and progress. BMJ Qual Saf. 2011;20(4):338–43. doi: 10.1136/bmjqs.2010.040964.

3. Okuyama A, Wagner C, Bijnen B. Speaking up for patient safety by hospital-based health care professionals: a literature review. BMC Health Serv Res. 2014;14:61–8. doi: 10.1186/1472-6963-14-61.

4. WHO: Global patient safety action plan 2021-2030. https://www.who.int/teams/integrated-health-services/patient-safety/policy/global-patient-safety-action-plan (2021). Accessed 01-03-2026.

5. WHO: Global patient safety action plan 2021–2030: implementation report 2024. (2024). Accessed 01-03-2026.

6. WHO: WHO global strategy on people-centred and integrated health services. (2015). Accessed 01-03-2026.

7. Santana MJ, Manalili K, Jolley RJ, Zelinsky S, Quan H, Lu M. How to practice person-centred care: A conceptual framework. Health Expect. 2018;21(2):429–40. doi: 10.1111/hex.12640.

8. WHO: People-centred care in health systems: improving quality and safety. (2022). Accessed 01-03-2026.

9. Ginsburg L, Castel E, Tregunno D and Norton PG. The H-PEPSS: an instrument to measure health professionals’ perceptions of patient safety competence at entry into practice. BMJ Quality & Safety. 2012;21(8):676–84. doi: 10.1136/bmjqs-2011-000601.

10. Alquwez N. Examining the influence of workplace incivility on nurses’ patient safety competence. Journal of Nursing Scholarship. 2020;52(3):292–300. doi: 10.1111/jnu.12553.

11. Zaitoun RA, Said NB and Tantillo LD. Clinical nurse competence and its effect on patient safety culture: a systematic review. BMC Nursing. 2023;22(1):173–82. doi: 10.1186/s12912-023-01305-w.

12. Ajzen I. The Theory of Planned Behavior, Organizational Behavior and Human Decision Processes. Organizational Behavior and Human Decision Processes. 1991;50(2):179–211. doi: 10.1016/0749-5978(91)90020-T.

13. Neal A, Griffin MA. A study of the lagged relationships among safety climate, safety motivation, safety behavior, and accidents at the individual and group levels. J Appl Psychol. 2006;91(4):946–53. doi: 10.1037/0021-9010.91.4.946.

14. Donabedian A. Evaluating the quality of medical care. 1966. Milbank Q. 2005;83(4):691-729. doi: 10.1111/j.1468-0009.2005.00397.x.

15. Reason J. Human error: models and management. BMJ. 2000;320(7237):768-70. doi: 10.1136/bmj.320.7237.768.

16. von Elm E, Altman DG, Egger M, Pocock SJ, Gøtzsche PC, Vandenbroucke JP. The Strengthening the Reporting of Observational Studies in Epidemiology (STROBE) statement: guidelines for reporting observational studies. J Clin Epidemiol. 2008;61(4):344–9. doi: 10.1016/j.jclinepi.2007.11.008.

17. Cohen J. Statistical Power Analysis for the Behavioral Sciences. 2nd ed ed. Hillsdale, NJ: Lawrence Erlbaum Associates; 1988.

18. Cochran WG. Sampling techniques, 3rd edition. New York: John Wiley & Sons; 1977.

19. WHO: Process of translation and adaptation of instruments. (2018). Accessed 01-03-2026.

20. Beaton DE, Bombardier C, Guillemin F, Ferraz MB. Guidelines for the process of cross-cultural adaptation of self-report measures. Spine (Phila Pa 1976). 2000;25(24):3186-91. doi: 10.1097/00007632-200012150-00014.

21. Edvardsson D, Fetherstonhaugh D, Nay R, Gibson S. Development and initial testing of the Person-centered Care Assessment Tool (P-CAT). Int Psychogeriatr. 2010;22(1):101–8. doi: 10.1017/s1041610209990688.

22. Park H-H, Kim S. A structural equation model of nurses’ patient safety management activities. Journal of Korean Academy of Nursing Administration. 2019;25(2):63–72. doi: 10.11111/jkana.2019.25.2.63.

23. WHO: Patient safety curriculum guide: multi-professional edition. (2011). Accessed 01-03-2026.

24. Hall KK, Shoemaker-Hunt S, Hoffman L, Richard S, Gall E, Schoyer E, et al. Making Healthcare Safer III: A Critical Analysis of Existing and Emerging Patient Safety Practices. Rockville (MD): Agency for Healthcare Research and Quality (US); 2020.

25. Sorra JS DN: Hospital Survey on Patient Safety Culture: User’s Guide. https://www.ahrq.gov/sops/surveys/hospital/index.html (2010). Accessed 01-03-2026.

26. Jacob Cohen PC, Stephen G. West, Leona S. Aiken. Applied Multiple Regression/Correlation Analysis for the Behavioral Sciences. 3rd ed ed. Mahwah, NJ: Lawrence Erlbaum Associates; 2003.

27. Preacher KJ, Hayes AF. Asymptotic and resampling strategies for assessing and comparing indirect effects in multiple mediator models. Behav Res Methods. 2008;40(3):879–91. doi: 10.3758/brm.40.3.879.

28. Thu NTH, Anh BTM, Ha NTT, Tien DNT, Giang PH, Nga TT, et al. Health staff perceptions of patient safety and associated factors in hospitals in Vietnam. Frontiers in Public Health. 2023;11. doi: 10.3389/fpubh.2023.1149667.

29. Tran TNH, Pham QT, Tran LH, Vu TA, Nguyen MT, Pham HT, et al. Comparison of perceptions about patient safety culture between physicians and nurses in public hospitals in Vietnam. Risk Management and Healthcare Policy. 2022:1695–704. doi: 10.2147/RMHP.S373249.

30. Atinafu D, Getaneh G, Setotaw G. Assessment of patient safety culture and associated factors among healthcare professionals in public hospitals of Bahir Dar City, Northwest Ethiopia: A mixed-methods study. PLoS One. 2024;19(11):e0313321. doi: 10.1371/journal.pone.0313321.

31. Boloré S, Sovet L, Guirimand N. Health professionals’ perceptions of patient safety competencies: psychometric properties of the French version of the H-PEPSS in France and Switzerland. BMC Medical Education. 2023;23(1):905–16. doi: 10.1186/s12909-023-04893-y.

32. Ha TTN, Thanh PQ, Huong TL, Anh VT, Tu NM, Tien PH, et al. Nurses’ perceptions about patient safety culture in public hospital in Vietnam. Appl Nurs Res. 2023;69:151650. doi: 10.1016/j.apnr.2022.151650.

33. Hafezi A, Babaii A, Aghaie B, Abbasinia M. The relationship between patient safety culture and patient safety competency with adverse events: a multicenter cross-sectional study. BMC Nursing. 2022;21(1):292–9. doi: 10.1186/s12912-022-01076-w.

34. Kim JJ, Mi JH. Effect of Patient Safety Culture and Patient Safety Competence on Safety Nursing Activity among Nurses working in Anesthetic and Recovery Rooms. J Korean Clin Nurs Res. 2020;26(2):164–74. doi: 10.22650/JKCNR.2020.26.2.164.

35. Park MN, Roh YS. Emergency department nurses’ reflective thinking and patient safety competency: The mediating effect of patient safety culture. Applied Nursing Research. 2024;80:151856. doi: 10.1016/j.apnr.2024.151856.

36. Tawfik DS, Adair KC, Palassof S, Sexton JB, Levoy E, Frankel A, et al. Leadership Behavior Associations with Domains of Safety Culture, Engagement, and Health Care Worker Well-Being. Jt Comm J Qual Patient Saf. 2023;49(3):156–65. doi: 10.1016/j.jcjq.2022.12.006.

